# Effects of BA.1/BA.2 subvariant, vaccination, and prior infection on infectiousness of SARS-CoV-2 Omicron infections

**DOI:** 10.1101/2022.03.02.22271771

**Authors:** Suelen H. Qassim, Hiam Chemaitelly, Houssein H. Ayoub, Sawsan AlMukdad, Patrick Tang, Mohammad R. Hasan, Hadi M. Yassine, Hebah A. Al-Khatib, Maria K. Smatti, Hanan F. Abdul-Rahim, Gheyath K. Nasrallah, Mohamed Ghaith Al-Kuwari, Abdullatif Al-Khal, Peter Coyle, Anvar Hassan Kaleeckal, Riyazuddin Mohammad Shaik, Ali Nizar Latif, Einas Al-Kuwari, Andrew Jeremijenko, Adeel A. Butt, Roberto Bertollini, Hamad Eid Al-Romaihi, Mohamed H. Al-Thani, Laith J. Abu-Raddad

## Abstract

**BACKGROUND:** Qatar experienced a large SARS-CoV-2 Omicron (B.1.1.529) wave that started on December 19, 2021 and peaked in mid-January, 2022. We investigated effects of Omicron subvariant (BA.1 and BA.2), previous vaccination, and prior infection on infectiousness of Omicron infections, between December 23, 2021 and February 20, 2022.

**METHODS:** Univariable and multivariable regression analyses were conducted to estimate the association between the RT-qPCR cycle threshold (Ct) value of PCR tests (a proxy for SARS-CoV-2 infectiousness) and each of the Omicron subvariants, mRNA vaccination, prior infection, reason for RT-qPCR testing, calendar week of RT-qPCR testing (to account for phases of the rapidly evolving Omicron wave), and demographic factors.

**RESULTS:** Compared to BA.1, BA.2 was associated with 3.53 fewer cycles (95% CI: 3.46-3.60), signifying higher infectiousness. Ct value decreased with time since second and third vaccinations. Ct values were highest for those who received their boosters in the month preceding the RT-qPCR test—0.86 cycles (95% CI: 0.72-1.00) higher than for unvaccinated persons. Ct value was 1.30 (95% CI: 1.20-1.39) cycles higher for those with a prior infection compared to those without prior infection, signifying lower infectiousness. Ct value declined gradually with age. Ct value was lowest for those who were tested because of symptoms and was highest for those who were tested for travel-related purposes. Ct value was lowest during the exponential-growth phase of the Omicron wave and was highest after the wave peaked and was declining.

**CONCLUSIONS:** The BA.2 subvariant appears substantially more infectious than the BA.1 subvariant. This may reflect higher viral load and/or longer duration of infection, thereby explaining the rapid expansion of this subvariant in Qatar.

## Introduction

Qatar experienced a large severe acute respiratory syndrome coronavirus 2 (SARS-CoV-2) Omicron (B.1.1.529)^1^ wave that started on December 19, 2021 and peaked in mid-January, 2022.^2-5^ We investigated effects of Omicron subvariant (BA.1 and BA.2), previous vaccination, and prior infection on infectiousness of Omicron infections, between December 23, 2021 and February 20, 2022. Incidence was initially dominated by BA.1, but within a few days, BA.2 predominated (Figure 1). Incidence of the Delta variant was minimal and no other variants were detected in viral genome sequencing and real-time reverse-transcription polymerase chain reaction (RT-qPCR) genotyping of randomly collected samples (Section S1 of Supplementary Appendix).

**Figure 1.**
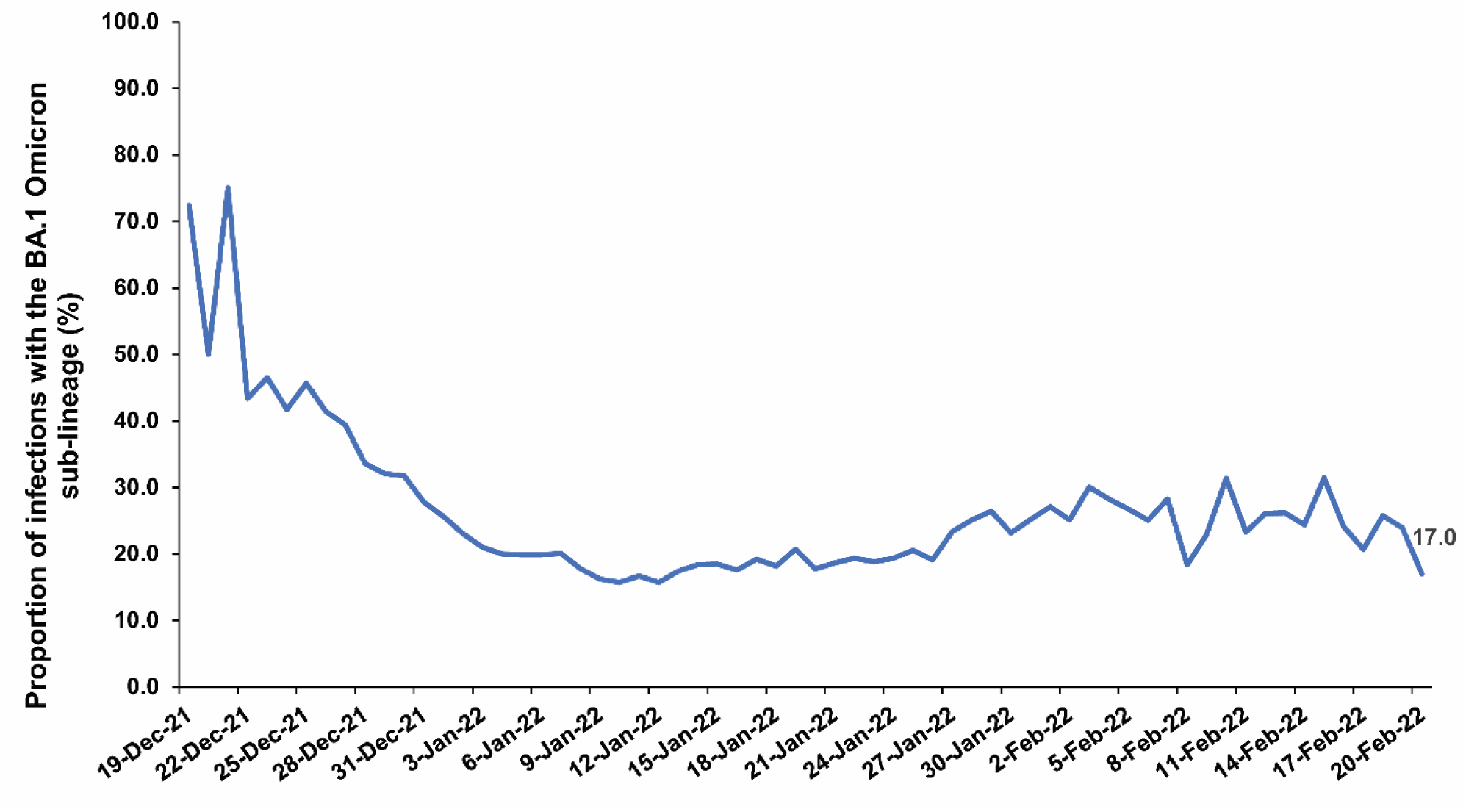
Proportion of BA.1 (versus BA.2) Omicron infections in the PCR-positive tests assessed using TaqPath COVID-19 Combo Kit during the study period.

## Methods

### Study population, data sources, and study design

The RT-qPCR cycle threshold (Ct) value is a measure of the inverse of viral load and correlates strongly with culturable virus;^6^ thus, it can be used as a proxy for SARS-CoV-2 infectiousness.^6-11^ We investigated several effects on the RT-qPCR Ct value of Omicron variant infections in the resident population of Qatar using a cross-sectional study design. These included: subvariant (BA.1 versus BA.2), mRNA (BNT162b2^12^ and mRNA-1273^13^) vaccination status, prior infection status, reason for RT-qPCR testing, study-period week of the RT-qPCR test (to account for the evolving phase of the rapid Omicron wave), and demographic factors including sex, age, and nationality.

The study population included all individuals with an RT-qPCR-confirmed SARS-CoV-2 infection in Qatar between December 23, 2021 and February 20, 2022. Coronavirus disease 2019 (COVID-19) laboratory testing, vaccination, clinical infection, and demographic data for this population were extracted from the national, federated SARS-CoV-2 databases, which include all RT-qPCR testing, reason for RT-qPCR testing, COVID-19 vaccinations, and related demographic details since the start of the pandemic. Further description of Qatar’s national COVID-19 databases can be found in previous publications.^11,14-17^

Every SARS-CoV-2 RT-qPCR test conducted in Qatar is classified based on the reason for testing (clinical symptoms, contact tracing, surveys or random testing campaigns, individual requests, routine healthcare testing, pre-travel, at port of entry, or other). RT-qPCR testing is performed at a mass scale.^15^ About 75% of those diagnosed over recent months were diagnosed not because of appearance of symptoms, but because of routine testing.^15^ Qatar has unusually young, diverse demographics, in that only 9% of its residents are ≥50 years of age, and 89% are expatriates from over 150 countries.^14,18^ Nearly all individuals were vaccinated in Qatar, however, vaccinations performed elsewhere were still recorded in the health system at the port of entry upon arrival to Qatar per country requirements.

Informed by the viral genome sequencing and the RT-qPCR genotyping (Section S1), a SARS-CoV-2 infection with the BA.1 subvariant was proxied as an S-gene “target failure” (SGTF) case using the TaqPath COVID-19 Combo Kit (Thermo Fisher Scientific, USA^19^) that tests for the S-gene and is affected by the del69/70 mutation in the S-gene.^20^ A SARS-CoV-2 infection with the BA.2 subvariant was proxied as a non-SGTF case using this TaqPath Kit. For ascertainment of subvariant status and standardization of RT-qPCR Ct values, we analyzed only the RT-qPCR-confirmed infections diagnosed with this TaqPath Kit.

For each individual, we selected only the first positive RT-qPCR-confirmed swab during the study period. We subsequently derived a summary measure for our primary outcome, the RT-qPCR Ct value, by averaging the Ct values of the N, ORF1ab, and S (if not an S-gene “target failure” case) genes. This average Ct value was used as the dependent variable in all analyses.

Both vaccination status and prior infection status were ascertained at the time of the RT-qPCR test. Vaccination status was defined factoring the number of administered vaccine doses and months elapsed since the last vaccine dose, with one month defined as 30 days. Only vaccination with BNT162b2^12^ and mRNA-1273^13^ vaccines were considered in the analyses, as these have been the vaccines of choice in the COVID-19 immunization program in Qatar.^21-23^ Rare occurrences of mixed vaccination regimens were excluded. Prior infection was defined as an RT-qPCR-positive test that occurred ≥90 days before the study RT-qPCR-positive test.^3,17,24-31^ An RT-qPCR-positive test that occurred <90 days prior to the study RT-qPCR-positive test was still factored in the analysis, but was not considered a prior infection. This is because this RT-qPCR-positive test and the study RT-qPCR-positive test may both reflect the same prolonged infection.^32-34^ A small number of RT-qPCR tests had no recorded Ct value and were thus excluded from the analysis, but these constituted only 0.1% of all RT-qPCR tests. Otherwise, data on the remaining study variables were complete.

### Oversight

Hamad Medical Corporation and Weill Cornell Medicine-Qatar Institutional Review Boards approved this retrospective study with waiver of informed consent. The study was reported following STROBE guidelines. The STROBE checklist is found in Table S1.

### Statistical analysis

Frequency distributions and measures of central tendency were used to describe the study population with respect to a priori determined factors. These included Omicron infection subvariant, vaccination status (factoring dose number and months since vaccination), prior infection status, reason for RT-qPCR testing, study-period week of the RT-qPCR test, and demographic factors, namely sex, age, and nationality. Differences between BA.1 and BA.2 infections were estimated using standardized mean differences (SMDs).

Association of each of these factors with Ct value was assessed using univariable linear regression analyses. Unadjusted β coefficients, 95% confidence intervals (CIs), and the F-test of overall covariate significance were reported. Adjusted β coefficients and associated 95% CIs and p-values were estimated using multivariable linear regression analyses that included all covariates in the model.

The 95% CIs were not adjusted for multiplicity. Two-sided p-value <0.05 indicated statistical significance. Interactions were not considered. Statistical analyses were conducted in STATA/SE version 16.^35^

## Results

Figure 2 shows the process of selecting the study population and Table 1 describes the study population characteristics. This was a national study involving 156,202 individuals infected with Omicron who are broadly representative of the population of Qatar.

**Figure 2.**
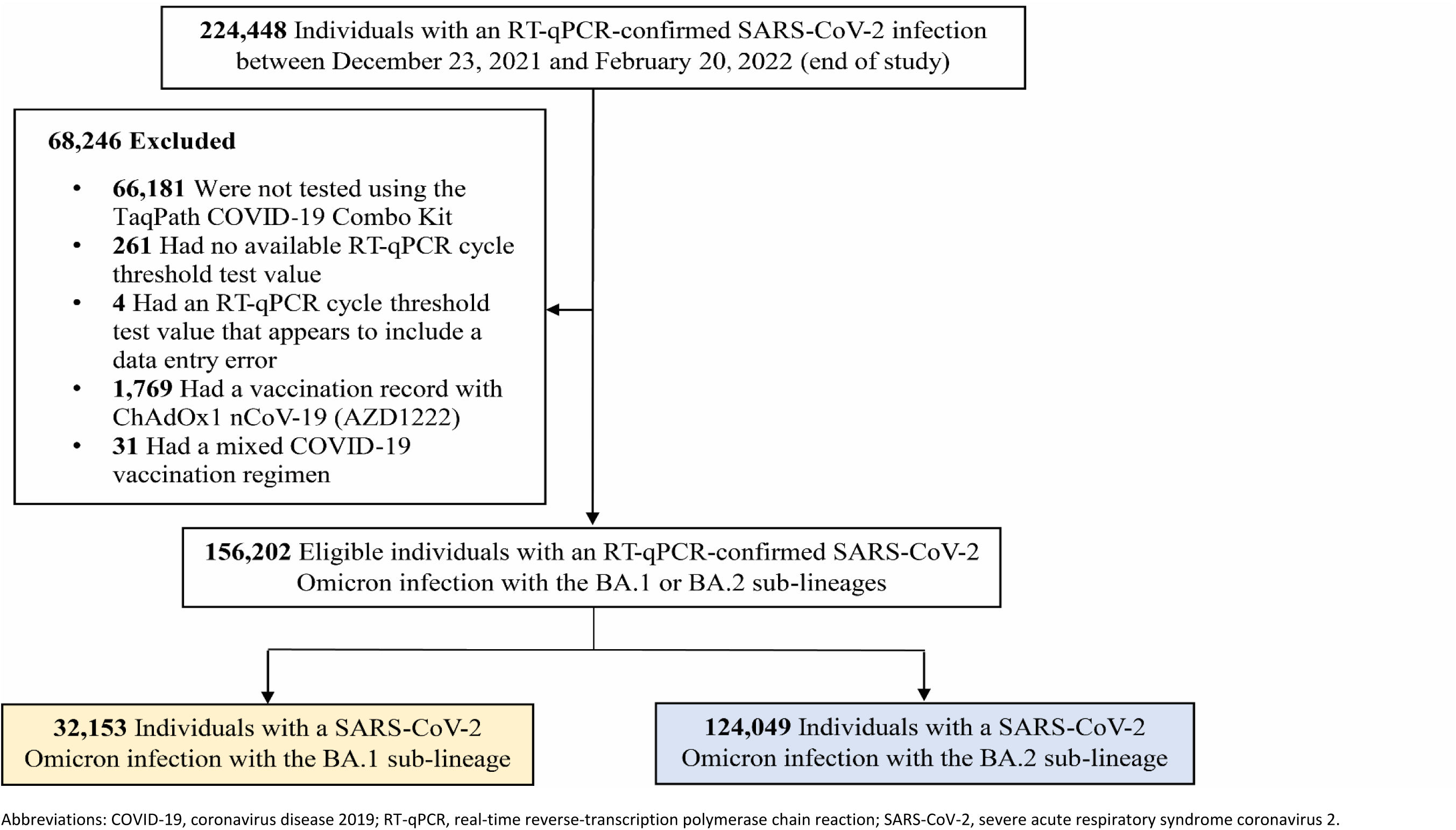
Flowchart describing the population selection process for investigating the infectiousness of SARS-CoV-2 Omicron variant infections.

**Table 1.**
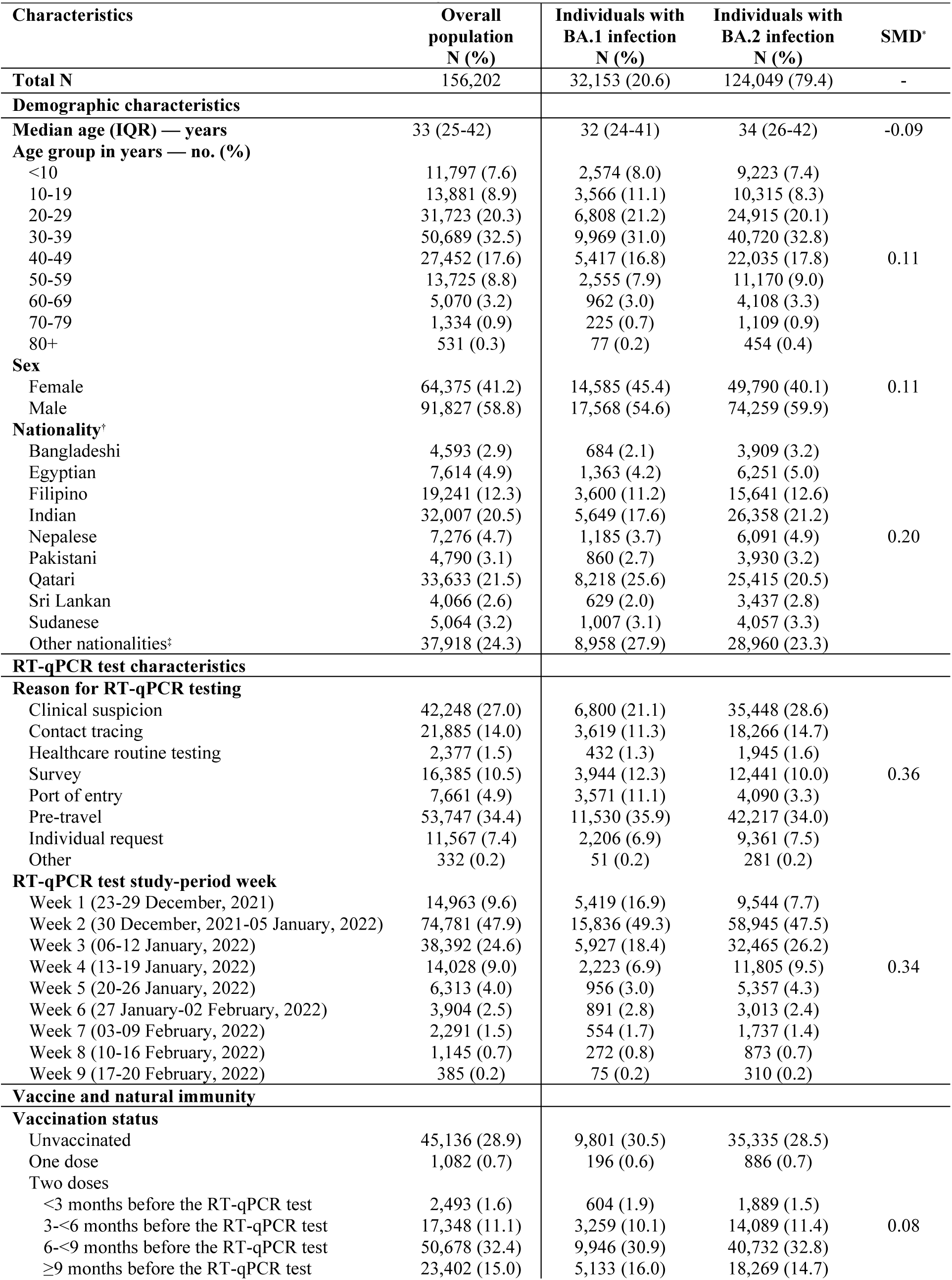
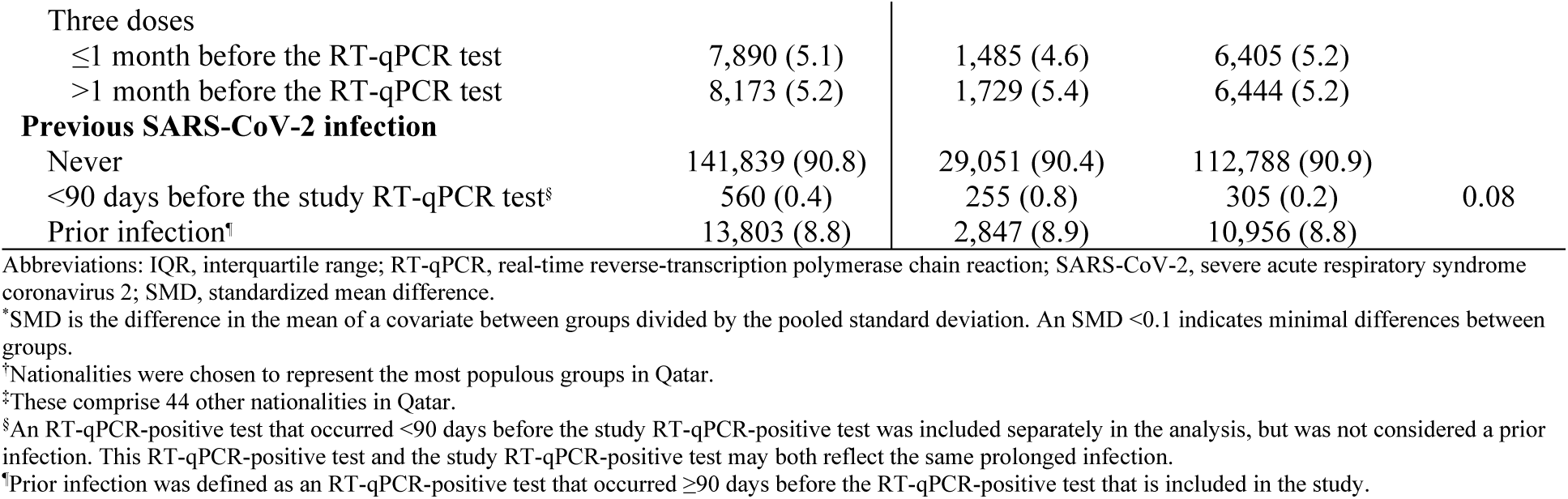
Characteristics of included individuals with SARS-CoV-2 Omicron infections between December 23, 2021 and February 20, 2022.

| Characteristics | Overall population<br>N (%) | Individuals with<br>BA.1 infection<br>N (%) | Individuals with<br>BA.2 infection<br>N (%) | SMD* |
| --- | --- | --- | --- | --- |
| **Total N** | 156,202 | 32,153 (20.6) | 124,049 (79.4) | - |
| **Demographic characteristics** |  |  |  |  |
| **Median age (IQR) — years** | 33 (25-42) | 32 (24-41) | 34 (26-42) | -0.09 |
| **Age group in years — no. (%)** |  |  |  |  |
| <10 | 11,797 (7.6) | 2,574 (8.0) | 9,223 (7.4) |  |
| 10-19 | 13,881 (8.9) | 3,566 (11.1) | 10,315 (8.3) |  |
| 20-29 | 31,723 (20.3) | 6,808 (21.2) | 24,915 (20.1) |  |
| 30-39 | 50,689 (32.5) | 9,969 (31.0) | 40,720 (32.8) |  |
| 40-49 | 27,452 (17.6) | 5,417 (16.8) | 22,035 (17.8) | 0.11 |
| 50-59 | 13,725 (8.8) | 2,555 (7.9) | 11,170 (9.0) |  |
| 60-69 | 5,070 (3.2) | 962 (3.0) | 4,108 (3.3) |  |
| 70-79 | 1,334 (0.9) | 225 (0.7) | 1,109 (0.9) |  |
| 80+ | 531 (0.3) | 77 (0.2) | 454 (0.4) |  |
| **Sex** |  |  |  |  |
| Female | 64,375 (41.2) | 14,585 (45.4) | 49,790 (40.1) | 0.11 |
| Male | 91,827 (58.8) | 17,568 (54.6) | 74,259 (59.9) |  |
| **Nationality†** |  |  |  |  |
| Bangladeshi | 4,593 (2.9) | 684 (2.1) | 3,909 (3.2) |  |
| Egyptian | 7,614 (4.9) | 1,363 (4.2) | 6,251 (5.0) |  |
| Filipino | 19,241 (12.3) | 3,600 (11.2) | 15,641 (12.6) |  |
| Indian | 32,007 (20.5) | 5,649 (17.6) | 26,358 (21.2) |  |
| Nepalese | 7,276 (4.7) | 1,185 (3.7) | 6,091 (4.9) | 0.20 |
| Pakistani | 4,790 (3.1) | 860 (2.7) | 3,930 (3.2) |  |
| Qatari | 33,633 (21.5) | 8,218 (25.6) | 25,415 (20.5) |  |
| Sri Lankan | 4,066 (2.6) | 629 (2.0) | 3,437 (2.8) |  |
| Sudanese | 5,064 (3.2) | 1,007 (3.1) | 4,057 (3.3) |  |
| Other nationalities‡ | 37,918 (24.3) | 8,958 (27.9) | 28,960 (23.3) |  |
| **RT-qPCR test characteristics** |  |  |  |  |
| **Reason for RT-qPCR testing** |  |  |  |  |
| Clinical suspicion | 42,248 (27.0) | 6,800 (21.1) | 35,448 (28.6) |  |
| Contact tracing | 21,885 (14.0) | 3,619 (11.3) | 18,266 (14.7) |  |
| Healthcare routine testing | 2,377 (1.5) | 432 (1.3) | 1,945 (1.6) |  |
| Survey | 16,385 (10.5) | 3,944 (12.3) | 12,441 (10.0) | 0.36 |
| Port of entry | 7,661 (4.9) | 3,571 (11.1) | 4,090 (3.3) |  |
| Pre-travel | 53,747 (34.4) | 11,530 (35.9) | 42,217 (34.0) |  |
| Individual request | 11,567 (7.4) | 2,206 (6.9) | 9,361 (7.5) |  |
| Other | 332 (0.2) | 51 (0.2) | 281 (0.2) |  |
| **RT-qPCR test study-period week** |  |  |  |  |
| Week 1 (23-29 December, 2021) | 14,963 (9.6) | 5,419 (16.9) | 9,544 (7.7) |  |
| Week 2 (30 December, 2021-05 January, 2022) | 74,781 (47.9) | 15,836 (49.3) | 58,945 (47.5) |  |
| Week 3 (06-12 January, 2022) | 38,392 (24.6) | 5,927 (18.4) | 32,465 (26.2) |  |
| Week 4 (13-19 January, 2022) | 14,028 (9.0) | 2,223 (6.9) | 11,805 (9.5) | 0.34 |
| Week 5 (20-26 January, 2022) | 6,313 (4.0) | 956 (3.0) | 5,357 (4.3) |  |
| Week 6 (27 January-02 February, 2022) | 3,904 (2.5) | 891 (2.8) | 3,013 (2.4) |  |
| Week 7 (03-09 February, 2022) | 2,291 (1.5) | 554 (1.7) | 1,737 (1.4) |  |
| Week 8 (10-16 February, 2022) | 1,145 (0.7) | 272 (0.8) | 873 (0.7) |  |
| Week 9 (17-20 February, 2022) | 385 (0.2) | 75 (0.2) | 310 (0.2) |  |
| **Vaccine and natural immunity** |  |  |  |  |
| **Vaccination status** |  |  |  |  |
| Unvaccinated | 45,136 (28.9) | 9,801 (30.5) | 35,335 (28.5) |  |
| One dose | 1,082 (0.7) | 196 (0.6) | 886 (0.7) |  |
| Two doses |  |  |  |  |
| <3 months before the RT-qPCR test | 2,493 (1.6) | 604 (1.9) | 1,889 (1.5) |  |
| 3-<6 months before the RT-qPCR test | 17,348 (11.1) | 3,259 (10.1) | 14,089 (11.4) | 0.08 |
| 6-<9 months before the RT-qPCR test | 50,678 (32.4) | 9,946 (30.9) | 40,732 (32.8) |  |
| ≥9 months before the RT-qPCR test | 23,402 (15.0) | 5,133 (16.0) | 18,269 (14.7) |  |
| Three doses |  |  |  |  |
| ≤1 month before the RT-qPCR test | 7,890 (5.1) | 1,485 (4.6) | 6,405 (5.2) |  |
| >1 month before the RT-qPCR test | 8,173 (5.2) | 1,729 (5.4) | 6,444 (5.2) |  |
| **Previous SARS-CoV-2 infection** |  |  |  |  |
| Never | 141,839 (90.8) | 29,051 (90.4) | 112,788 (90.9) |  |
| <90 days before the study RT-qPCR test<sup>§</sup> | 560 (0.4) | 255 (0.8) | 305 (0.2) | 0.08 |
| Prior infection<sup>¶</sup> | 13,803 (8.8) | 2,847 (8.9) | 10,956 (8.8) |  |

Abbreviations: IQR, interquartile range; RT-qPCR, real-time reverse-transcription polymerase chain reaction; SARS-CoV-2, severe acute respiratory syndrome coronavirus 2; SMD, standardized mean difference.
<sup>†</sup>SMD is the difference in the mean of a covariate between groups divided by the pooled standard deviation. An SMD <0.1 indicates minimal differences between groups.
<sup>‡</sup>Nationalities were chosen to represent the most populous groups in Qatar.
<sup>§</sup>These comprise 44 other nationalities in Qatar.
<sup>§</sup>An RT-qPCR-positive test that occurred <90 days before the study RT-qPCR-positive test was included separately in the analysis, but was not considered a prior infection. This RT-qPCR-positive test and the study RT-qPCR-positive test may both reflect the same prolonged infection.
<sup>¶</sup>Prior infection was defined as an RT-qPCR-positive test that occurred ≥90 days before the RT-qPCR-positive test that is included in the study.

Compared to BA.1, BA.2 was associated with 3.53 fewer cycles (95% CI: 3.46-3.60), signifying higher infectiousness (Table 2). Ct value decreased with time since second and third vaccinations, mirroring the established pattern of waning vaccine effectiveness.^4^ Ct values were highest for those who received their boosters in the month preceding the RT-qPCR test—0.86 cycles (95% CI: 0.72-1.00) higher than for unvaccinated persons. Ct value was 1.30 (95% CI: 1.20-1.39) cycles higher for those with a prior infection compared to those without prior infection, signifying lower infectiousness.

**Table 2.**
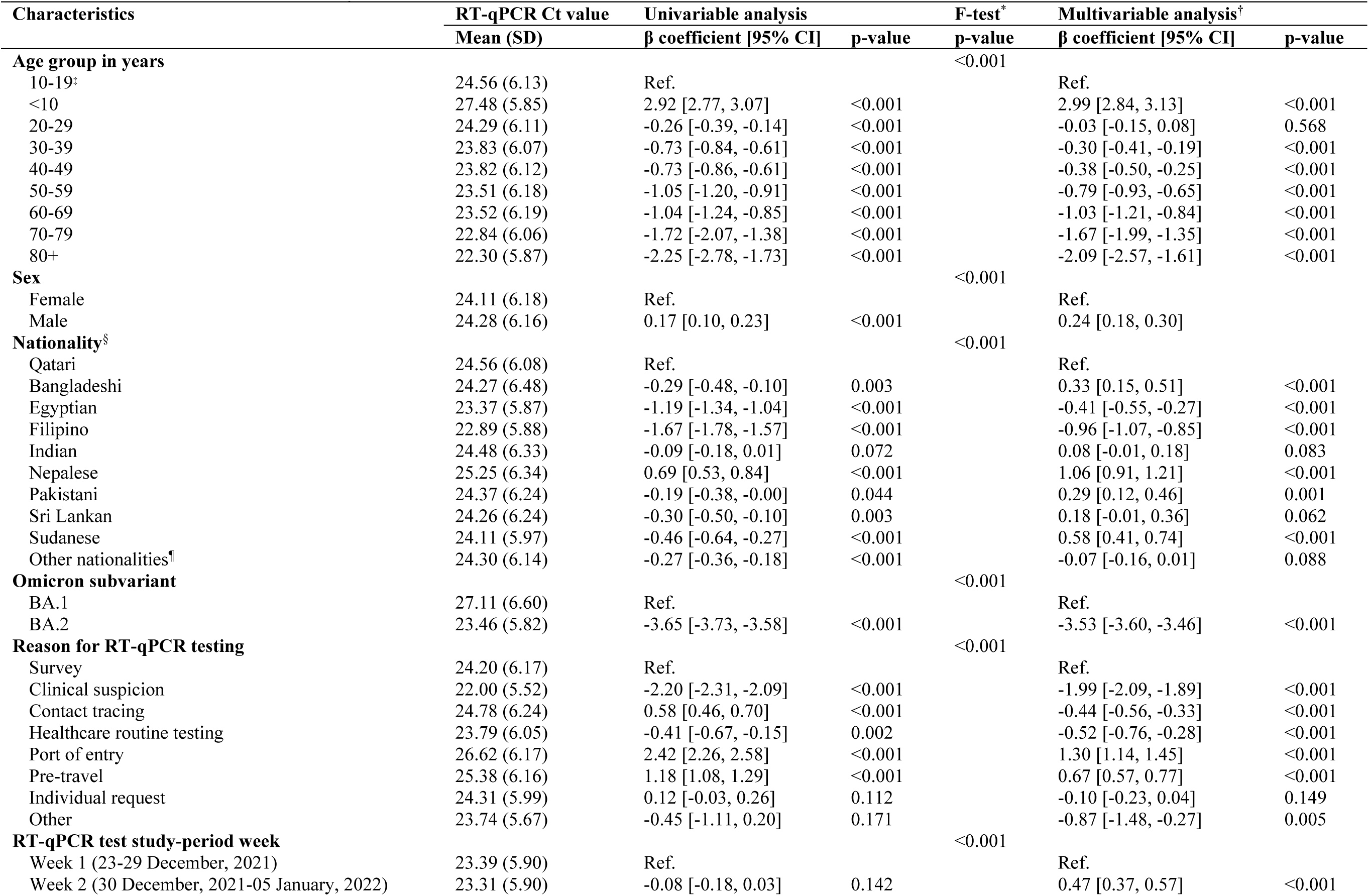
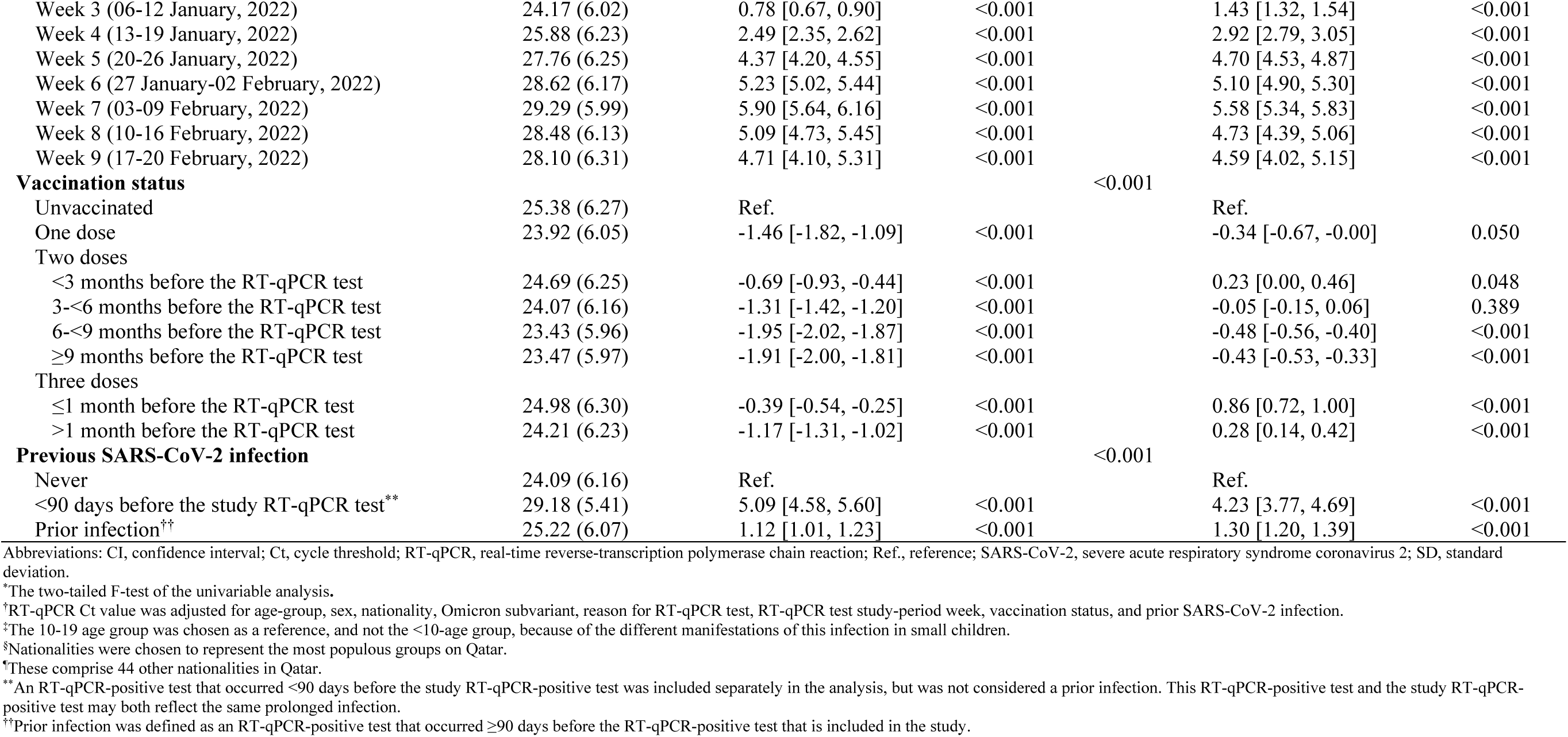
Associations with RT-qPCR Ct value among 156,202 individuals with SARS-CoV-2 Omicron infection between December 23, 2021 and February 20, 2022.

Ct value declined gradually with age (Table 2), perhaps reflecting slower virus clearance with aging. There were differences in Ct value by sex and nationality, but these may reflect different test-seeking behaviors for different socio-economic groups in Qatar’s diverse population, or differences in the rates of prior undocumented infection by nationality.^14,36-39^ Ct value was lowest for those who were tested because of symptoms and was highest for those who were tested for travel-related purposes. Ct value was lowest during the exponential-growth phase of the Omicron wave, as a large proportion of infections were recent, and was highest after the wave peaked and was declining, as a small proportion of infections were recent. Stratified analyses for each of BA.1 (Table 3) and BA.2 (Table 4) showed similar findings.

**Table 3.** Associations with RT-qPCR Ct value among 32,153 individuals with SARS-CoV-2 Omicron BA.1 subvariant infection.

| Characteristics | RT-qPCR Ct value | Univariable analysis |  | F-test* | Multivariable analysis† |  |
| --- | --- | --- | --- | --- | --- | --- |
|  | Mean (SD) | β coefficient [95% CI] | p-value | p-value | β coefficient [95% CI] | p-value |
| **Age group in years** |  |  |  | <0.001 |  |  |
| 10-19‡ | 26.96 (6.40) | Ref. |  |  | Ref. |  |
| <10 | 30.17 (5.51) | 3.21 [2.88, 3.54] | <0.001 |  | 3.09 [2.77, 3.42] | <0.001 |
| 20-29 | 27.20 (6.52) | 0.24 [-0.03, 0.50] | 0.082 |  | 0.09 [-0.16, 0.35] | 0.469 |
| 30-39 | 26.76 (6.69) | -0.20 [-0.45, 0.05] | 0.113 |  | -0.23 [-0.48, 0.02] | 0.069 |
| 40-49 | 26.69 (6.66) | -0.27 [-0.55, 0.01] | 0.055 |  | -0.37 [-0.64, -0.10] | 0.008 |
| 50-59 | 26.69 (6.76) | -0.27 [-0.61, 0.06] | 0.105 |  | -0.56 [-0.89, -0.24] | 0.001 |
| 60-69 | 26.59 (6.68) | -0.37 [-0.83, 0.10] | 0.123 |  | -0.93 [-1.37, -0.49] | <0.001 |
| 70-79 | 25.69 (6.95) | -1.27 [-2.15, -0.39] | 0.005 |  | -1.74 [-2.56, -0.92] | <0.001 |
| 80+ | 25.14 (7.10) | -1.82 [-3.30, -0.35] | 0.015 |  | -2.41 [-3.77, -1.04] | 0.001 |
| **Sex** |  |  |  | <0.001 |  |  |
| Female | 26.67 (6.57) | Ref. |  |  | Ref. |  |
| Male | 27.48 (6.60) | 0.81 [0.67, 0.96] | <0.001 |  | 0.31 [0.16, 0.45] | <0.001 |
| **Nationality§** |  |  |  | <0.001 |  |  |
| Qatari | 26.89 (6.34) | Ref. |  |  | Ref. |  |
| Bangladeshi | 28.95 (6.87) | 2.06 [1.55, 2.57] | <0.001 |  | 1.49 [1.01, 1.98] | <0.001 |
| Egyptian | 26.22 (6.45) | -0.66 [-1.04, -0.29] | 0.001 |  | -0.49 [-0.84, -0.14] | 0.006 |
| Filipino | 25.21 (6.69) | -1.67 [-1.93, -1.42] | <0.001 |  | -1.22 [-1.47, -0.96] | <0.001 |
| Indian | 28.08 (6.76) | 1.20 [0.98, 1.42] | <0.001 |  | 0.47 [0.24, 0.69] | <0.001 |
| Nepalese | 29.42 (6.55) | 2.54 [2.14, 2.93] | <0.001 |  | 1.88 [1.49, 2.26] | <0.001 |
| Pakistani | 27.60 (6.54) | 0.72 [0.26, 1.18] | 0.002 |  | 0.00 [-0.43, 0.43] | 0.999 |
| Sri Lankan | 28.38 (6.76) | 1.49 [0.96, 2.02] | <0.001 |  | 0.92 [0.42, 1.42] | <0.001 |
| Sudanese | 26.85 (6.47) | -0.03 [-0.46, 0.40] | 0.890 |  | 0.38 [-0.02, 0.78] | 0.065 |
| Other nationalities¶ | 27.05 (6.47) | 0.17 [-0.03, 0.36] | 0.092 |  | -0.07 [-0.26, 0.12] | 0.466 |
| **Reason for RT-qPCR testing** |  |  |  | <0.001 |  |  |
| Survey | 26.69 (6.53) | Ref. |  |  | Ref. |  |
| Clinical suspicion | 24.43 (6.43) | -2.26 [-2.51, -2.01] | <0.001 |  | -2.20 [-2.44, -1.96] | <0.001 |
| Contact tracing | 28.10 (6.78) | 1.42 [1.13, 1.71] | <0.001 |  | -0.03 [-0.30, 0.25] | 0.854 |
| Healthcare routine testing | 27.28 (6.53) | 0.59 [-0.05, 1.23] | 0.069 |  | 0.02 [-0.58, 0.62] | 0.943 |
| Port of entry | 27.65 (6.09) | 0.96 [0.67, 1.25] | <0.001 |  | 0.52 [0.24, 0.79] | <0.001 |
| Pre-travel | 28.31 (6.37) | 1.63 [1.39, 1.86] | <0.001 |  | 0.61 [0.38, 0.83] | <0.001 |
| Individual request | 27.37 (6.50) | 0.68 [0.34, 1.01] | <0.001 |  | 0.09 [-0.22, 0.41] | 0.569 |
| Other | 25.86 (6.93) | -0.83 [-2.61, 0.95] | 0.360 |  | -1.32 [-2.98, 0.35] | 0.121 |
| **RT-qPCR test study-period week** |  |  |  | <0.001 |  |  |
| Week 1 (23-29 December, 2021) | 25.81 (6.39) | Ref. |  |  | Ref. |  |
| Week 2 (30 December, 2021-05 January, 2022) | 25.96 (6.40) | 0.14 [-0.05, 0.34] | 0.148 |  | 0.17 [-0.02, 0.35] | 0.083 |
| Week 3 (06-12 January, 2022) | 27.76 (6.50) | 1.94 [1.71, 2.18] | <0.001 |  | 1.69 [1.46, 1.91] | <0.001 |
| Week 4 (13-19 January, 2022) | 30.32 (5.99) | 4.50 [4.19, 4.81] | <0.001 |  | 3.97 [3.66, 4.27] | <0.001 |
| Week 5 (20-26 January, 2022) | 32.08 (5.36) | 6.27 [5.84, 6.70] | <0.001 |  | 5.65 [5.23, 6.07] | <0.001 |
| Week 6 (27 January-02 February, 2022) | 32.32 (5.04) | 6.51 [6.06, 6.95] | <0.001 |  | 5.84 [5.40, 6.27] | <0.001 |
| Week 7 (03-09 February, 2022) | 33.11 (4.36) | 7.29 [6.74, 7.84] | <0.001 |  | 6.70 [6.16, 7.23] | <0.001 |
| Week 8 (10-16 February, 2022) | 32.04 (5.15) | 6.23 [5.46, 6.99] | <0.001 |  | 5.48 [4.74, 6.22] | <0.001 |
| Week 9 (17-20 February, 2022) | 31.29 (5.53) | 5.48 [4.04, 6.91] | <0.001 |  | 5.10 [3.72, 6.48] | <0.001 |
| **Vaccination status** |  |  |  | <0.001 |  |  |
| Unvaccinated | 28.36 (6.40) | Ref. |  |  | Ref. |  |
| One dose | 27.24 (6.63) | -1.12 [-2.04, -0.19] | 0.018 |  | -0.46 [-1.32, 0.39] | 0.288 |
| Two doses |  |  |  |  |  |  |
| <3 months before the RT-qPCR test | 27.19 (6.39) | -1.17 [-1.71, -0.63] | <0.001 |  | 0.08 [-0.42, 0.58] | 0.758 |
| 3-<6 months before the RT-qPCR test | 26.98 (6.72) | -1.38 [-1.63, -1.12] | <0.001 |  | -0.17 [-0.43, 0.08] | 0.180 |
| 6-<9 months before the RT-qPCR test | 26.23 (6.57) | -2.13 [-2.31, -1.94] | <0.001 |  | -0.66 [-0.85, -0.47] | <0.001 |
| ≥9 months before the RT-qPCR test | 26.07 (6.54) | -2.29 [-2.51, -2.07] | <0.001 |  | -0.67 [-0.90, -0.45] | <0.001 |
| Three doses |  |  |  |  |  |  |
| ≤1 month before the RT-qPCR test | 28.42 (6.59) | 0.06 [-0.29, 0.42] | 0.724 |  | 0.96 [0.62, 1.31] | <0.001 |
| >1 month before the RT-qPCR test | 27.29 (6.60) | -1.07 [-1.40, -0.74] | <0.001 |  | 0.29 [-0.03, 0.62] | 0.077 |
| **Previous SARS-CoV-2 infection** |  |  |  | <0.001 |  |  |
| Never | 26.97 (6.62) | Ref. |  |  | Ref. |  |
| <90 days before the study RT-qPCR test** | 30.87 (4.92) | 3.90 [3.09, 4.71] | <0.001 |  | 4.67 [3.93, 5.42] | <0.001 |
| Prior infection†† | 28.24 (6.32) | 1.27 [1.02, 1.53] | <0.001 |  | 1.61 [1.37, 1.84] | <0.001 |

Abbreviations: CI, confidence interval; Ct, cycle threshold; RT-qPCR, real-time reverse-transcription polymerase chain reaction; Ref., reference; SARS-CoV-2, severe acute respiratory syndrome coronavirus 2; SD, standard deviation.
\*The two-tailed F-test of the univariable analysis.
†RT-qPCR Ct value was adjusted for age-group, sex, nationality, Omicron subvariant, reason for RT-qPCR test, RT-qPCR test study-period week, vaccination status, and prior SARS-CoV-2 infection.
‡The 10-19 age group was chosen as a reference, and not the <10-age group, because of the different manifestations of this infection in small children.
§Nationalities were chosen to represent the most populous groups on Qatar.
\*These comprise 44 other nationalities in Qatar.
\*\*An RT-qPCR-positive test that occurred <90 days before the study RT-qPCR-positive test was included separately in the analysis, but was not considered a prior infection. This RT-qPCR-positive test and the study RT-qPCR-positive test may both reflect the same prolonged infection.
††Prior infection was defined as an RT-qPCR-positive test that occurred ≥90 days before the RT-qPCR-positive test that is included in the study.

**Table 4.** Associations with RT-qPCR Ct value among 124,049 individuals with SARS-CoV-2 Omicron BA.2 subvariant infection.

| Characteristics | RT-qPCR Ct value | Univariable analysis |  | F-test* | Multivariable analysis† |  |
| --- | --- | --- | --- | --- | --- | --- |
|  | Mean (SD) | β coefficient [95% CI] | p-value | p-value | β coefficient [95% CI] | p-value |
| **Age group in years** |  |  |  | <0.001 |  |  |
| 10-19‡ | 23.73 (5.81) | Ref. |  |  | Ref. |  |
| <10 | 26.73 (5.72) | 3.00 [2.84, 3.16] | <0.001 |  | 2.95 [2.80, 3.11] | <0.001 |
| 20-29 | 23.50 (5.74) | -0.23 [-0.36, -0.10] | 0.001 |  | -0.09 [-0.22, 0.04] | 0.160 |
| 30-39 | 23.12 (5.68) | -0.61 [-0.74, -0.49] | <0.001 |  | -0.34 [-0.47, -0.22] | <0.001 |
| 40-49 | 23.12 (5.77) | -0.61 [-0.74, -0.47] | <0.001 |  | -0.40 [-0.54, -0.27] | <0.001 |
| 50-59 | 22.78 (5.80) | -0.95 [-1.11, -0.80] | <0.001 |  | -0.85 [-1.01, -0.70] | <0.001 |
| 60-69 | 22.79 (5.84) | -0.93 [-1.14, -0.73] | <0.001 |  | -1.07 [-1.27, -0.87] | <0.001 |
| 70-79 | 22.26 (5.70) | -1.47 [-1.83, -1.12] | <0.001 |  | -1.68 [-2.02, -1.34] | <0.001 |
| 80+ | 21.82 (5.51) | -1.90 [-2.44, -1.37] | <0.001 |  | -2.06 [-2.57, -1.55] | <0.001 |
| **Sex** |  |  |  | <0.001 |  |  |
| Female | 23.37 (5.85) | Ref. |  |  | Ref. |  |
| Male | 23.52 (5.80) | 0.16 [0.09, 0.22] | <0.001 |  | 0.20 [0.13, 0.26] | <0.001 |
| **Nationality§** |  |  |  | <0.001 |  |  |
| Qatari | 23.81 (5.80) | Ref. |  |  | Ref. |  |
| Bangladeshi | 23.46 (6.04) | -0.36 [-0.55, -0.16] | <0.001 |  | 0.06 [-0.13, 0.25] | 0.512 |
| Egyptian | 22.75 (5.54) | -1.06 [-1.22, -0.90] | <0.001 |  | -0.45 [-0.60, -0.29] | <0.001 |
| Filipino | 22.35 (5.54) | -1.46 [-1.57, -1.34] | <0.001 |  | -0.96 [-1.08, -0.85] | <0.001 |
| Indian | 23.70 (5.96) | -0.11 [-0.21, -0.01] | 0.033 |  | -0.08 [-0.18, 0.03] | 0.150 |
| Nepalese | 24.44 (5.97) | 0.63 [0.47, 0.79] | <0.001 |  | 0.81 [0.65, 0.97] | <0.001 |
| Pakistani | 23.67 (5.95) | -0.15 [-0.34, 0.05] | 0.139 |  | 0.29 [0.11, 0.48] | 0.002 |
| Sri Lankan | 23.51 (5.83) | -0.30 [-0.51, -0.10] | 0.004 |  | -0.03 [-0.23, 0.16] | 0.731 |
| Sudanese | 23.43 (5.64) | -0.39 [-0.58, -0.20] | <0.001 |  | 0.57 [0.39, 0.75] | <0.001 |
| Other nationalities¶ | 23.44 (5.77) | -0.37 [-0.47, -0.27] | <0.001 |  | -0.12 [-0.21, -0.02] | 0.015 |
| **Reason for RT-qPCR testing** |  |  |  | <0.001 |  |  |
| Survey | 23.41 (5.84) | Ref. |  |  | Ref. |  |
| Clinical suspicion | 21.53 (5.20) | -1.88 [-2.00, -1.76] | <0.001 |  | -1.95 [-2.06, -1.84] | <0.001 |
| Contact tracing | 24.12 (5.91) | 0.71 [0.58, 0.84] | <0.001 |  | -0.50 [-0.63, -0.38] | <0.001 |
| Healthcare routine testing | 23.01 (5.66) | -0.40 [-0.67, -0.13] | 0.004 |  | -0.63 [-0.89, -0.37] | <0.001 |
| Port of entry | 25.72 (6.09) | 2.31 [2.11, 2.51] | <0.001 |  | 1.96 [1.77, 2.15] | <0.001 |
| Pre-travel | 24.58 (5.86) | 1.17 [1.06, 1.29] | <0.001 |  | 0.68 [0.57, 0.79] | <0.001 |
| Individual request | 23.60 (5.63) | 0.19 [0.03, 0.34] | 0.017 |  | -0.15 [-0.29, -0.00] | 0.048 |
| Other | 23.36 (5.34) | -0.05 [-0.72, 0.62] | 0.886 |  | -0.80 [-1.44, -0.16] | 0.014 |
| **RT-qPCR test study-period week** |  |  |  | <0.001 |  |  |
| Week 1 (23-29 December, 2021) | 22.02 (5.11) | Ref. |  |  | Ref. |  |
| Week 2 (30 December, 2021-05 January, 2022) | 22.60 (5.55) | 0.59 [0.46, 0.71] | <0.001 |  | 0.51 [0.39, 0.62] | <0.001 |
| Week 3 (06-12 January, 2022) | 23.52 (5.69) | 1.50 [1.38, 1.63] | <0.001 |  | 1.32 [1.19, 1.45] | <0.001 |
| Week 4 (13-19 January, 2022) | 25.04 (5.91) | 3.03 [2.87, 3.18] | <0.001 |  | 2.64 [2.49, 2.79] | <0.001 |
| Week 5 (20-26 January, 2022) | 26.99 (6.09) | 4.98 [4.79, 5.17] | <0.001 |  | 4.45 [4.27, 4.64] | <0.001 |
| Week 6 (27 January-02 February, 2022) | 27.53 (6.04) | 5.51 [5.28, 5.74] | <0.001 |  | 4.78 [4.56, 5.01] | <0.001 |
| Week 7 (03-09 February, 2022) | 28.07 (5.93) | 6.06 [5.77, 6.35] | <0.001 |  | 5.15 [4.87, 5.43] | <0.001 |
| Week 8 (10-16 February, 2022) | 27.37 (5.98) | 5.36 [4.97, 5.75] | <0.001 |  | 4.41 [4.03, 4.79] | <0.001 |
| Week 9 (17-20 February, 2022) | 27.33 (6.25) | 5.31 [4.67, 5.95] | <0.001 |  | 4.40 [3.78, 5.01] | <0.001 |
| **Vaccination status** |  |  |  | <0.001 |  |  |
| Unvaccinated | 24.55 (5.97) | Ref. |  |  | Ref. |  |
| One dose | 23.18 (5.66) | -1.36 [-1.75, -0.98] | <0.001 |  | -0.32 [-0.68, 0.04] | 0.083 |
| Two doses |  |  |  |  |  |  |
| <3 months before the RT-qPCR test | 23.89 (5.99) | -0.66 [-0.93, -0.39] | <0.001 |  | 0.31 [0.05, 0.56] | 0.017 |
| 3-<6 months before the RT-qPCR test | 23.39 (5.82) | -1.16 [-1.27, -1.04] | <0.001 |  | 0.01 [-0.10, 0.12] | 0.827 |
| 6-<9 months before the RT-qPCR test | 22.75 (5.59) | -1.80 [-1.88, -1.72] | <0.001 |  | -0.42 [-0.51, -0.33] | <0.001 |
| ≥9 months before the RT-qPCR test | 22.74 (5.59) | -1.81 [-1.91, -1.71] | <0.001 |  | -0.34 [-0.45, -0.24] | <0.001 |
| Three doses |  |  |  |  |  |  |
| ≤1 month before the RT-qPCR test | 24.19 (5.96) | -0.36 [-0.52, -0.21] | <0.001 |  | 0.84 [0.69, 0.99] | <0.001 |
| >1 month before the RT-qPCR test | 23.38 (5.86) | -1.17 [-1.32, -1.01] | <0.001 |  | 0.30 [0.15, 0.46] | <0.001 |
| **Previous SARS-CoV-2 infection** |  |  |  | <0.001 |  |  |
| Never | 23.35 (5.82) | Ref. |  |  | Ref. |  |
| <90 days before the study RT-qPCR test** | 27.78 (5.40) | 4.42 [3.77, 5.07] | <0.001 |  | 3.95 [3.34, 4.56] | <0.001 |
| Prior infection†† | 24.43 (5.75) | 1.08 [0.96, 1.19] | <0.001 |  | 1.23 [1.12, 1.33] | <0.001 |

Abbreviations: CI, confidence interval; Ct, cycle threshold; RT-qPCR, real-time reverse-transcription polymerase chain reaction; Ref., reference; SARS-CoV-2, severe acute respiratory syndrome coronavirus 2; SD, standard deviation.
\*The two-tailed F-test of the univariable analysis.
†RT-qPCR Ct value was adjusted for age-group, sex, nationality, Omicron subvariant, reason for RT-qPCR test, RT-qPCR test study-period week, vaccination status, and prior SARS-CoV-2 infection.
‡The 10-19 age group was chosen as a reference, and not the <10-age group, because of the different manifestations of this infection in small children.
§Nationalities were chosen to represent the most populous groups on Qatar.
¶These comprise 44 other nationalities in Qatar.
\*\*An RT-qPCR-positive test that occurred <90 days before the study RT-qPCR-positive test was included separately in the analysis, but was not considered a prior infection. This RT-qPCR-positive test and the study RT-qPCR-positive test may both reflect the same prolonged infection.
††Prior infection was defined as an RT-qPCR-positive test that occurred ≥90 days before the RT-qPCR-positive test that is included in the study.

## Discussion

The BA.2 subvariant appears substantially more infectious than the BA.1 subvariant, consistent with recent findings of a household study from Denmark.^40^ This may reflect higher viral load and/or longer duration of infection, thereby explaining the rapid expansion of this subvariant in Qatar (Figure 1). Natural immunity from previous infection and strength of vaccine immunity correlate with less infectious breakthrough infections, as observed for earlier SARS-CoV-2 variants.^11^ Symptomatic infection and older age are associated with higher infectiousness.

### Limitations and caveats

A small number of RT-qPCR tests had no available Ct value and were thus excluded from the analysis, but these constituted only 0.1% of all RT-qPCR tests. The study was implemented on documented RT-qPCR-confirmed infections, but other infections may have occurred but were never documented. It is possible that infections in those with prior infection or those vaccinated are less likely to be diagnosed, perhaps because of minimal or no symptoms. Nevertheless, RT-qPCR testing in Qatar is done at a mass scale, where a significant proportion of the population is being tested every week.^15^ The majority of infections are identified not because of symptoms, but because of routine testing for other reasons (Table 1).^15^ The date of symptom onset was not available for symptomatic cases. Therefore, an analysis factoring the duration between symptom onset and RT-qPCR test was not possible.

The study population consists of mostly working-age adults and thus the results may not necessarily be generalizable to other population groups, such as the elderly. The analyses controlled for sex, age, and nationality but it was not possible to control for other factors, such as comorbidities or socio-economic factors, as data on these factors were not available. Of note that the number of individuals with severe chronic conditions is small in Qatar’s young population.^14,41^ The national list of vaccine prioritization included only 19,800 individuals of all age groups with serious co-morbid conditions to be prioritized in the first phase of vaccine roll-out.^15^ Factoring nationality in the analyses may have (partially) controlled socio-economic differences/occupational risk, in consideration of the association between nationality and occupation in Qatar.^14,36-39^

BA.1 and BA.2 ascertainment was based on proxy criteria; presence or absence of SGTF using the TaqPath Kit, but this method of ascertainment is well established not only for Omicron subvariants, but also for other variants such as Alpha.^20,30,42^ Some Omicron infections may have been misclassified Delta infections, but this is not likely, as Delta incidence was limited during the study (Section S1).

Time since vaccination was associated with lower Ct value, mirroring the established pattern of waning vaccine effectiveness.^4^ Unexpectedly, however, the Ct value for those who had their second dose >6 months earlier was lower than that among unvaccinated persons. With the high vaccine coverage in Qatar (exceeding 85%), the unvaccinated group may be different in other uncontrolled attributes from the vaccinated group. For example, those unvaccinated may have chosen not to receive the vaccine because of undocumented prior infection and thus are not truly immune naïve.

## Data Availability

The dataset of this study is a property of the Qatar Ministry of Public Health that was provided to the researchers through a restricted-access agreement that prevents sharing the dataset with a third party or publicly. Future access to this dataset can be considered through a direct application for data access to Her Excellency the Minister of Public Health (https://www.moph.gov.qa/english/Pages/default.aspx). Aggregate data are available within the manuscript and its Supplementary information.

## Acknowledgements

We acknowledge the many dedicated individuals at Hamad Medical Corporation, the Ministry of Public Health, the Primary Health Care Corporation, the Qatar Biobank, Sidra Medicine, and Weill Cornell Medicine – Qatar for their diligent efforts and contributions to make this study possible.

The authors are grateful for support from the Biomedical Research Program and the Biostatistics, Epidemiology, and Biomathematics Research Core, both at Weill Cornell Medicine-Qatar, as well as for support provided by the Ministry of Public Health, Hamad Medical Corporation, and Sidra Medicine. The authors are also grateful for the Qatar Genome Programme and Qatar University Biomedical Research Center for institutional support for the reagents needed for the viral genome sequencing. Statements made herein are solely the responsibility of the authors. The funders of the study had no role in study design, data collection, data analysis, data interpretation, or writing of the article.

## Author contributions

SHQ co-designed the study, performed the statistical analyses, and co-wrote the first draft of the article. HC co-designed the study, supported the statistical analyses, and co-wrote the first draft of the article. LJA conceived and co-designed the study, led the statistical analyses, and co-wrote the first draft of the article. PT and MRH conducted the multiplex, RT-qPCR variant screening and viral genome sequencing. HY, HAK, and MS conducted viral genome sequencing. All authors contributed to data collection and acquisition, database development, discussion and interpretation of the results, and to the writing of the manuscript. All authors have read and approved the final manuscript.

## Supplementary Appendix

### Section S1. Laboratory methods and variant ascertainment

#### Real-time reverse-transcription polymerase chain reaction testing

Nasopharyngeal and/or oropharyngeal swabs were collected for polymerase chain reaction (PCR) testing and placed in Universal Transport Medium (UTM). Aliquots of UTM were: 1) extracted on KingFisher Flex (Thermo Fisher Scientific, USA), MGISP-960 (MGI, China), or ExiPrep 96 Lite (Bioneer, South Korea) followed by testing with real-time reverse-transcription PCR (RT-qPCR) using TaqPath COVID-19 Combo Kits (Thermo Fisher Scientific, USA) on an ABI 7500 FAST (Thermo Fisher Scientific, USA); 2) tested directly on the Cepheid GeneXpert system using the Xpert Xpress SARS-CoV-2 (Cepheid, USA); or 3) loaded directly into a Roche cobas 6800 system and assayed with the cobas SARS-CoV-2 Test (Roche, Switzerland). The first assay targets the viral S, N, and ORF1ab gene regions. The second targets the viral N and E-gene regions, and the third targets the ORF1ab and E-gene regions.

All PCR testing was conducted at the Hamad Medical Corporation Central Laboratory or Sidra Medicine Laboratory, following standardized protocols.

#### Classification of infections by variant type

Surveillance for severe acute respiratory syndrome coronavirus 2 (SARS-CoV-2) variants in Qatar is mainly based on viral genome sequencing and multiplex RT-qPCR variant screening^1^ of random positive clinical samples,^2-7^ complemented by deep sequencing of wastewater samples.^4,8^

A total of 315 random SARS-CoV-2-positive specimens collected between December 19, 2021 and January 22, 2022 were viral whole-genome sequenced on a Nanopore GridION sequencing device. Of these, 300 (95.2%) were confirmed as Omicron (B.1.1.529)^9^ infections and 15 (4.8%) as Delta (B.1.617.2)^9^ infections.^4,10,11^ Of 286 Omicron infections with confirmed subvariant status, 68 (23.8%) were BA.1 cases and 218 (76.2%) were BA.2 cases. No Delta case was detected in sequencing after January 8, 2022, nor were other variants.

Additionally, a total of 1,315 random SARS-CoV-2-positive specimens collected between December 22, 2021 and January 1, 2022 were RT-qPCR genotyped. The RT-qPCR genotyping identified 1 B.1.617.2-like Delta case, 366 BA.1-like Omicron cases, 898 BA.2-like Omicron cases, and 50 were undetermined cases where the genotype could not be assigned.

The accuracy of the RT-qPCR genotyping was verified against either Sanger sequencing of the receptor-binding domain (RBD) of SARS-CoV-2 surface glycoprotein (S) gene, or by viral whole-genome sequencing on a Nanopore GridION sequencing device. From 147 random SARS-CoV-2-positive specimens all collected in December of 2021, RT-qPCR genotyping was able to assign a genotype in 129 samples. The agreement between RT-qPCR genotyping and sequencing was 100% for Delta (n=82), 100% for Omicron BA.1 (n=18), and 93% for Omicron BA.2 (27 of 29 were correctly assigned to BA.2 and remaining 2 specimens genotyped as BA.2 were B.1.617.2 by sequencing). Of the remaining 18 specimens: 10 failed PCR amplification and sequencing, 8 could not be assigned a genotype by RT-qPCR (4 of 8 were B.1.617.2 by sequencing, and the remaining 4 failed sequencing). All the variant RT-qPCR genotyping was conducted at the Sidra Medicine Laboratory following standardized protocols.

The large Omicron-wave exponential-growth phase in Qatar started on December 19, 2021 and peaked in mid-January, 2022.^4,10-13^ The study duration coincided with the intense Omicron wave where Delta incidence was limited. Accordingly, any PCR-positive test during the study duration, between December 23, 2021 and February 20, 2022, was assumed to be an Omicron infection. Of note that the study duration started on December 23, 2021, and not on December 19, 2021, to minimize the occurrence of residual Delta incidence during the first few days of the Omicron wave.

Informed by the viral genome sequencing and the RT-qPCR genotyping, a SARS-CoV-2 infection with the BA.1 subvariant was proxied as an S-gene “target failure” (SGTF) case using the TaqPath COVID-19 Combo Kit (Thermo Fisher Scientific, USA^14^) that tests for the S-gene and is affected by the del69/70 mutation in the S-gene.^15^ A SARS-CoV-2 infection with the BA.2 subvariant was proxied as a non-SGTF case using this TaqPath Kit. For ascertainment of subvariant status and standardization of RT-qPCR cycle threshold values, we analyzed only the RT-qPCR-confirmed infections diagnosed with this TaqPath Kit.

**Table S1.** STROBE checklist for cross-sectional studies.

|  | Item No | Recommendations | Main text page No |
| --- | --- | --- | --- |
| Title and abstract | 1 | (a) Indicate the study’s design with a commonly used term in the title or the abstract | Methods (‘Study population, data sources, and study design’, paragraph 1) |
|  |  | (b) Provide in the abstract an informative and balanced summary of what was done and what was found | Abstract |
| Introduction |  |  |  |
| Background/rationale | 2 | Explain the scientific background and rationale for the investigation being reported | Introduction |
| Objectives | 3 | State specific objectives, including any prespecified hypotheses | Methods (‘Study population, data sources, and study design’) |
| Methods |  |  |  |
| Study design | 4 | Present key elements of study design early in the paper | Methods (‘Study population, data sources, and study design’) |
| Setting | 5 | Describe the setting, locations, and relevant dates, including periods of recruitment, exposure, follow-up, and data collection | Methods (‘Study population, data sources, and study design’) and Section S1 (‘Classification of infections by variant type’) of Supplementary Appendix |
| Participants | 6 | (a) Give the eligibility criteria, and the sources and methods of selection of participants | Methods (‘Study population, data sources, and study design’) & Figure 2 |
| Variables | 7 | Clearly define all outcomes, exposures, predictors, potential confounders, and effect modifiers. Give diagnostic criteria, if applicable | Methods (‘Study population, data sources, and study design’ & ‘Statistical analysis’) & Section S1 (‘Classification of infections by variant type’) of Supplementary Appendix |
| Data sources/<br>measurement | 8 | For each variable of interest, give sources of data and details of methods of assessment (measurement). Describe comparability of assessment methods if there is more than one group | Methods (‘Study population, data sources, and study design’), Table 1, & Section S1 of Supplementary Appendix |
| Bias | 9 | Describe any efforts to address potential sources of bias | Methods (‘Study population, data sources, and study design’ & ‘Statistical analysis’) |
| Study size | 10 | Explain how the study size was arrived at | Methods (‘Study population, data sources, and study design’) & Figure 2 |
| Quantitative variables | 11 | Explain how quantitative variables were handled in the analyses. If applicable, describe which groupings were chosen and why | Methods (‘Study population, data sources, and study design’ & ‘Statistical analysis’), & Tables 1-4 |
| Statistical methods | 12 | (a) Describe all statistical methods, including those used to control for confounding | Methods (‘Statistical analysis’) |
|  |  | (b) Describe any methods used to examine subgroups and interactions | Methods (‘Study population, data sources, and study design’ & ‘Statistical analysis’), & Tables 1, 3 & 4 |
|  |  | (c) Explain how missing data were addressed | Methods (‘Study population, data sources, and study design’) & Figure 2 |
|  |  | (d) If applicable, describe analytical methods taking account of sampling strategy | NA |
|  |  | (e) Describe any sensitivity analyses | NA |
| Results |  |  |  |
| Participants | 13 | (a) Report numbers of individuals at each stage of study—eg numbers potentially eligible, examined for eligibility, confirmed eligible, included in the study, completing follow-up, and analysed<br>(b) Give reasons for non-participation at each stage<br>(c) Consider use of a flow diagram | Figure 2 |
| Descriptive data | 14 | (a) Give characteristics of study participants (eg demographic, clinical, social) and information on exposures and potential confounders | Table 1 |

|  |  | (b) Indicate number of participants with missing data for each variable of interest | Methods ('Study population, data sources, and study design') & Figure 2 |
| --- | --- | --- | --- |
| Outcome data | 15 | Report numbers of outcome events or summary measures | Results & Table 1 |
| Main results | 16 | (a) Give unadjusted estimates and, if applicable, confounder-adjusted estimates and their precision (eg, 95% confidence interval). Make clear which confounders were adjusted for and why they were included | Results & Table 2 |
|  |  | (b) Report category boundaries when continuous variables were categorized | Tables 1-2 |
|  |  | (c) If relevant, consider translating estimates of relative risk into absolute risk for a meaningful time period | NA |
| Other analyses | 17 | Report other analyses done—eg analyses of subgroups and interactions, and sensitivity analyses | Results & Tables 3-4 |
| **Discussion** |  |  |  |
| Key results | 18 | Summarise key results with reference to study objectives | Discussion, paragraph 1 |
| Limitations | 19 | Discuss limitations of the study, taking into account sources of potential bias or imprecision. Discuss both direction and magnitude of any potential bias | Discussion ('Limitations and caveats') |
| Interpretation | 20 | Give a cautious overall interpretation of results considering objectives, limitations, multiplicity of analyses, results from similar studies, and other relevant evidence | Discussion, paragraph 1 |
| Generalisability | 21 | Discuss the generalisability (external validity) of the study results | Discussion ('Limitations and caveats') |
| **Other information** |  |  |  |
| Funding | 22 | Give the source of funding and the role of the funders for the present study and, if applicable, for the original study on which the present article is based | Acknowledgements |

Abbreviations: NA: not applicable.

